# Epidemiology and Nomogram Development for Chronic Eosinophilic Leukemia, Not Otherwise Specified (CEL-NOS): Insights from the SEER Database

**DOI:** 10.1101/2023.09.29.23296356

**Authors:** Fan Wang

**Author notes:** Correspondence: Dr. Fan Wang, Department of Hematology, Tongji Hospital, Tongji Medical College, Huazhong University of Science and Technology, 1095 Jiefang Avenue, Wuhan, Hubei 430030, China.

## Abstract

**Background:** Chronic Eosinophilic Leukemia, Not Otherwise Specified (CEL-NOS), a rare and intricate hematological disorder characterized by uncontrolled eosinophilic proliferation, presents clinical challenges owing to its infrequency. This study aimed to investigate the epidemiology and develop a prognostic nomogram for CEL-NOS patients.

**Methods:** Utilizing the Surveillance, Epidemiology and End Results (SEER) database, CEL-NOS cases diagnosed between 2001 and 2020 were analyzed for incidence rates, clinical profiles, and survival outcomes. Patients were randomly divided into training and validation cohorts (7:3 ratio). LASSO regression analysis and Cox regression analysis were performed to screen the prognostic factors for overall survival. A nomogram was then constructed and validated to predict the 3- and 5-year overall survival probability of CEL-NOS patients by incorporating these factors.

**Results:** The incidence rate of CEL-NOS was very low, with an average of 0.033 per 100,000 person-years from 2001 to 2020. The incidence rate significantly increased with age and was higher in males than females. The mean age at diagnosis was 57 years. Prognostic analysis identified advanced age, specific marital statuses, and secondary CEL-NOS as independent and adverse predictors of overall survival (OS). To facilitate personalized prognostication, a nomogram was developed incorporating these factors, demonstrating good calibration and discrimination. Risk stratification using the nomogram effectively differentiated patients into low- and high-risk groups.

**Conclusions:** This study enhances our understanding of CEL-NOS, offering novel insights into its epidemiology, demographics, and prognostic determinants, while providing a possible prognostication tool for clinical use. However, further research is warranted to elucidate molecular mechanisms and optimize therapeutic strategies for CEL-NOS.

## Introduction

Chronic eosinophilic leukemia, not otherwise specified (CEL-NOS), is an infrequent and challenging subtype of BCR-ABL negative myeloproliferative neoplasm (MPN) that primarily targets eosinophils-a crucial component of the body’s immune response and allergy regulation^1,2^. Characterized by specific genetic attributes, CEL-NOS is set apart by the absence of PDGFRA, PDGFRB, or FGFR1 rearrangements and PCM1-JAK2, ETV6-JAK2, or BCR-JAK2 fusion genes^2,3^. This distinctive neoplasm is marked by the persistent proliferation of eosinophilic precursors, resulting in pronounced eosinophilia (> 1.5 x 10^9^/L) in both peripheral blood and bone marrow^2^. Differentiating CEL-NOS from other eosinophilic disorders, often driven by specific genetic alterations or secondary factors, is crucial^4^. While the precise etiology of CEL-NOS remains elusive, its diagnosis hinges on the exclusion of alternative causes of eosinophilia. Two pivotal factors distinguish CEL-NOS from hypereosinophilic syndromes (HES): (a) the presence of clonality among eosinophils, as indicated by cytogenetic or molecular abnormalities; and (b) the existence of excess myeloblasts (<20%) in peripheral blood (>2%) or bone marrow (>5%)^4,5^. Additionally, rigorous exclusion of non-myeloid malignancies and myeloid neoplasms with eosinophilia, such as acute myeloid leukemia (AML) with inv16, chronic myeloid leukemia, myeloproliferative neoplasms, and myelodysplastic syndrome, is imperative^6^.

The clinical presentation of CEL-NOS is highly variable and contingent on the degree and duration of eosinophilia, the extent of organ involvement, and the risk of progression to AML^7^. Patients may experience a spectrum of symptoms, including fatigue, weakness, weight loss, fever, night sweats, splenomegaly, hepatomegaly, lymphadenopathy, and so on^7,8^. These complications can be life-threatening and require swift recognition and management^8^. Moreover, some patients with CEL-NOS may progress to AML, a more aggressive hematological malignancy with a less favorable prognosis^7^. As a result, patients with CEL-NOS necessitate regular follow-up and monitoring to assess their disease status and response to therapy.

Managing CEL-NOS is a complex endeavor, typically necessitating a multidisciplinary approach^8^. Therapeutic strategies aim to reduce eosinophilia and prevent or reverse organ damage^2^. However, no standardized therapy exists for CEL-NOS, and treatment options are often limited in their efficacy^2,8^. The choice of therapy is influenced by the presence of specific genetic abnormalities that may predict a patient’s response to certain drugs^9^. For instance, patients with FIP1L1-PDGFRA rearrangements or other PDGFRA mutations may benefit from imatinib, a tyrosine kinase inhibitor that targets aberrant signaling pathways^9,10^. Nevertheless, most CEL-NOS patients lack these mutations and may exhibit resistance or intolerance to imatinib^10^. For such cases, alternative treatment options encompass corticosteroids, hydroxyurea, interferon-alpha, mepolizumab and chemotherapy^2,11-13^. However, these therapies have variable efficacy, significant toxicity profiles, and may not alter the natural progression of the disease, underscoring the critical need for improved therapeutic strategies^7^. Furthermore, most CEL-NOS patients are ineligible for allogeneic stem cell transplantation, a possible curative option, due to their advanced age and significant comorbidities^13^. Participation in clinical trials may offer hope for novel and more effective therapeutic options for this challenging condition.

Data associated with the prognosis of CEL-NOS is limited. Prognosis in CEL-NOS may dependent on factors such as the degree and duration of eosinophilia, the extent and reversibility of organ damage, the presence of cytogenetic or molecular abnormalities, and the response to therapy^14^. Nevertheless, some patients experience a more aggressive clinical course marked by life-threatening complications or progression to AML^7^. According to a study of a small cohort with 10 patients, the survival durations in CEL-NOS vary, ranging from 2.2 months to 10 years (median survival was 22.2 months)^7^. Adverse prognostic markers encompass abnormal karyotypes, increased blast counts, thrombocytopenia, bone marrow fibrosis, atypical megakaryocytes, resistance, or intolerance to imatinib, and transformation to AML^7,14^.

Owing to the intricacies of diagnosis and the rarity of CEL-NOS, comprehensive investigations among patients meeting the 2016 World Health Organization (WHO) criteria for CEL-NOS have been notably limited. The purpose of this study is to investigate the incidence and identify the factors affecting the survival of CEL-NOS patients based on a population-based study using the national Cancer Institute’s Surveillance, Epidemiology and End Results (SEER) database. Moreover, we endeavor to construct a nomogram to predict the prognosis of patients with CEL-NOS.

## Materials and Methods

### Study Population and Data acquisition

Data for this study were sourced from the Surveillance, Epidemiology, and End Results (SEER) Program (https://seer.cancer.gov/), maintained by the National Cancer Institute (NCI). The data were collected using SEER*Stat software version 8.4.1 (https://seer.cancer.gov/seerstat/, accessed on August 1, 2023). The SEER 17 database [Incidence-SEER Research Data, 17 registries, Nov 2022 Sub (2000-2020)] was utilized to extract data of disease incidence by using the “rate session”. Patients diagnosed with CEL-NOS between 2001 and 2020 were selected from the “Incidence-SEER Research Plus Data, 17 Registries, Nov 2022 Sub (2000-2020)” database using the “case listing session”. Only cases with known age (censored at age 89 years) and malignant behavior were included. The inclusion criteria were as follows: the International Classification of Diseases for Oncology (ICD-O-3) histologic code (9964/3). The exclusion criteria were as follows: (1) the diagnosis confirmation was “unknown”; (2) the patient’s survival time was 0 or unknown; (3) age below 20 years old. Ultimately, 487 patients with CEL-NOS were included in the final cohort. Ethical approval was not required as SEER data are publicly available and anonymized, precluding patient reidentification. A flowchart depicting the selection process is presented in Figure 1.

**Figure 1.**
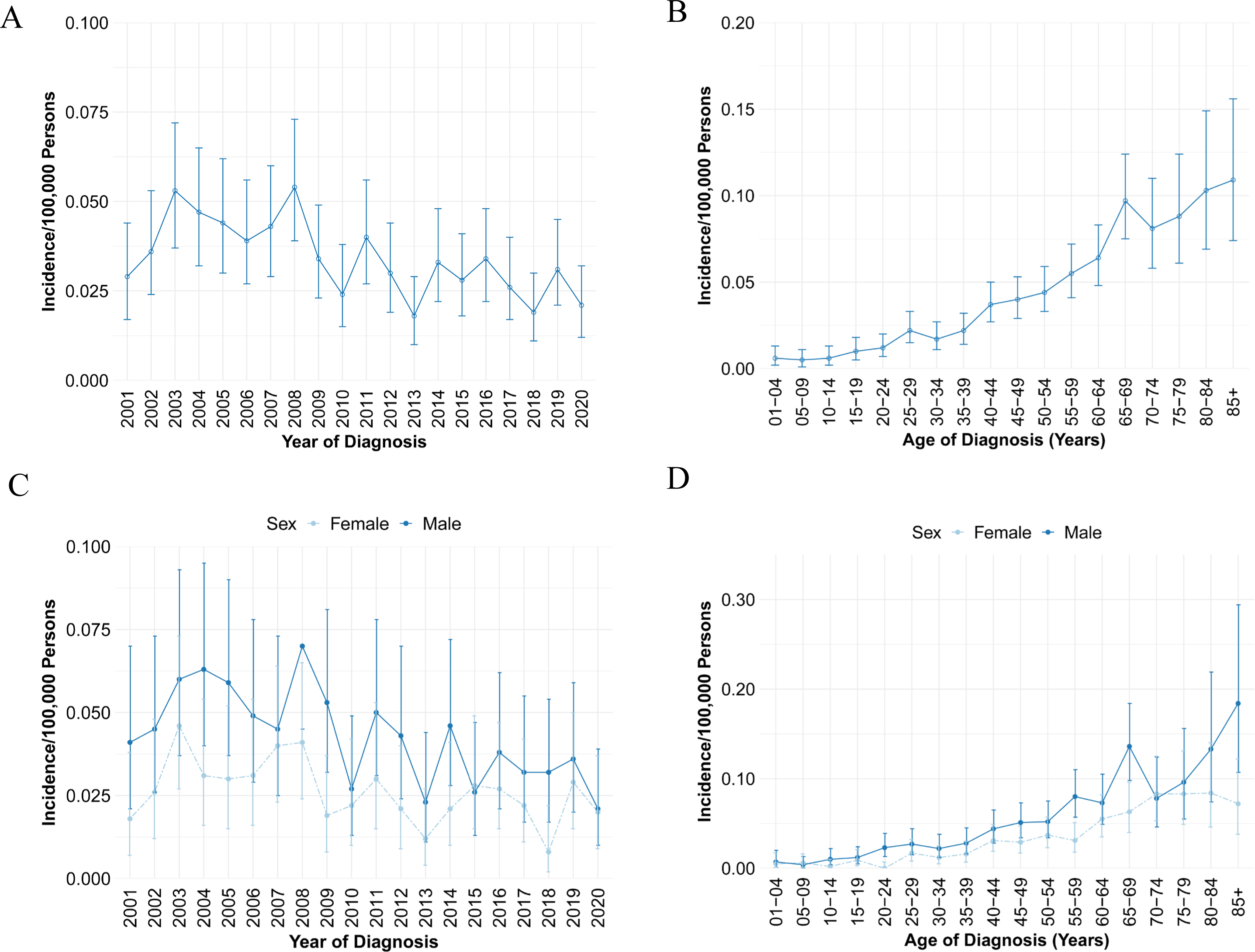
Age-adjusted incidence of CEL-NOS from 2001 to 2020 in SEER database. (A) Annual age-adjusted incidence of CEL-NOS; (B) Age-adjusted incidence of CEL-NOS based on age of diagnosis; (C) Annual age-adjusted incidence of CEL-NOS in male and female populations, respectively; (D) Age-adjusted incidence of CEL-NOS based on age of diagnosis in male and female patients, respectively. CEL-NOS, chronic eosinophilic leukemia, not otherwise specified.

### Definition of Variables

The analysis encompassed an array of variables, including age, sex, race, marital status, year of diagnosis, primary site, vital status, survival months, cause of death (COD) to site recode, cause-specific death classification, cause of death to site, sequence number, first malignant primary indicator, total number of in situ/malignant tumors for the patient, type of reporting source, diagnostic confirmation, surgery of the primary site, chemotherapy recode, and radiation recode. Age was dichotomized into <60 years and 60+ years, referencing the age at diagnosis of CEL-NOS. Race categories included African American, White, and Other (comprising “Asian/Pacific Islander,” “American Indian/Alaska Native,” and “Unknown”). Marital status was classified as married, single, or other (encompassing “divorced,” “separated,” “widowed,” “unmarried or domestic partner,” and “unknown”). COD information was derived from the “COD to site recode” field. In the SEER database, CEL-NOS-related death was defined as “dead (attributed to this cancer diagnosis),” while CEL-NOS-unrelated death was defined as death “dead (attributable to causes other than this cancer diagnosis).” Diagnosis years were categorized into “2001-2005,” “2006-2010,” “2011-2015,” and “2016-2020.” Overall survival (OS) time was calculated from the date of diagnosis to death or last follow-up.

### Statistical analysis

All statistical analyses were conducted using the R program language (version 4.2.1; R Foundation for Statistical Computing, Vienna, Austria). Patients were randomly allocated to training and validation groups (7:3 ratio), with baseline characteristics compared using Student’s *t*-test and *chi*-square test for continuous and categorical data, respectively. Prognostic risk factors for CEL-NOS were identified through least absolute shrinkage and selection operator (LASSO) regression, followed by univariate and multivariate Cox proportional regression analyses to determine independent prognostic factors. Hazard ratios (HR) and 95% confidence intervals (95% CIs) were reported. The Cox proportional hazards regression model was assessed for proportionality assumptions, with no violations detected. Variables with *P*_J<_J0.05 in the multivariate model were considered significant. Nomograms predicting 3- and 5-year overall survival (OS) probabilities were constructed based on independent prognostic factors. Time-dependent receiver operating characteristic (ROC) curves and corresponding area under the curve (AUC) values assessed discrimination. Calibration curves and decision curve analysis (DCA) nomograms were generated to evaluate performance. Patients from both cohorts were classified into high- and low-risk groups using median nomogram points, with Kaplan-Meier analysis employed for OS estimation and log-rank tests for group comparisons. A two-sided *P* value < 0.05 indicated statistical significance.

## Results

### Incidence of CEL-NOS

The analysis of data from the SEER database showed that the age-adjusted incidence rate (AIR) of CEL-NOS from 2001 to 2020 [age adjusted to the 2000 US Standard Population (19 age groups - Census P25-1130)] was 0.033 per 100,000 person-years (95% CI, 0.031-0.036). The annual AIR of CEL-NOS was presented in Figure 1A. Notably, the peak AIR was documented in 2008 with 0.054 per 100,000 person-years (95% CI, 0.039-0.073). Moreover, it is notable that the AIR of CEL-NOS increased with age (Figure 1B). Compared with the AIR of patients <60 years old (0.024/100,000 person-years), the incidence rate ratio (IRR) for the 60+ age group (AIR 0.087/100,000 person years) was 3.65 (95% CI, 3.07-4.34, *P* < 0.0001), indicating statistical significance. Investigation into gender differences revealed that the AIR of male (0.042/100,000 person-years) was significantly higher than that of female (0.025/100,000 person-years, IRR 1.66, 95% CI, 1.39-1.98, *P* < 0.0001, Figure 1C, D).

### Baseline Characteristics of CEL-NOS Patients

As depicted in Figure 2, 487 patients were finally identified as CEL-NOS in the SEER 17 registry, Nov 2022 Sub (2000-2020) from January 2001 to December 2020. The primary site was bone marrow (*n* = 487, 100%). For all patients in the study cohort, 61.2% were male, 1.6 folds that of female (*n*=189, 38.8%; Table 1). The average age at diagnosis was 57.0 ± 17.0 years (Range: 20-89), with 53.4% under the age of 60 (< 60) and 46.6% aged 60 or older (60+). Most CEL-NOS patients were white (73.9%), African American and Other (including Asian/Pacific Islander and American Indian/Alaska Native) occupied 15.6% and 10.5%, respectively. At diagnosis, 57.3% of patients were married, 24.6% had other marital statuses (such as divorced, separated, widowed, unmarried or domestic partner), and 18.1% were single who had never been married. For all CEL-NOS cases in this study, 89.7% were primary CEL-NOS and 10.3% were secondary CEL-NOS that were secondary to other primary malignancies. About 41.3% were treated with chemotherapy. At the time of last follow-up, 284 (58.3%) patients were alive; 42 (8.6%) deaths were attributable to CEL-NOS, 45 (9.2%) patients died of heart diseases, and an additional 116 (23.8%) patients died due to other causes such as diabetes mellitus, cerebrovascular diseases, septicemia, and so on. For all those variables, there were no statistical difference between the training cohort and validation cohort (*P* > 0.05). The comparison of epidemiologic characteristics was summarized in Table 1.

**Figure 2.**
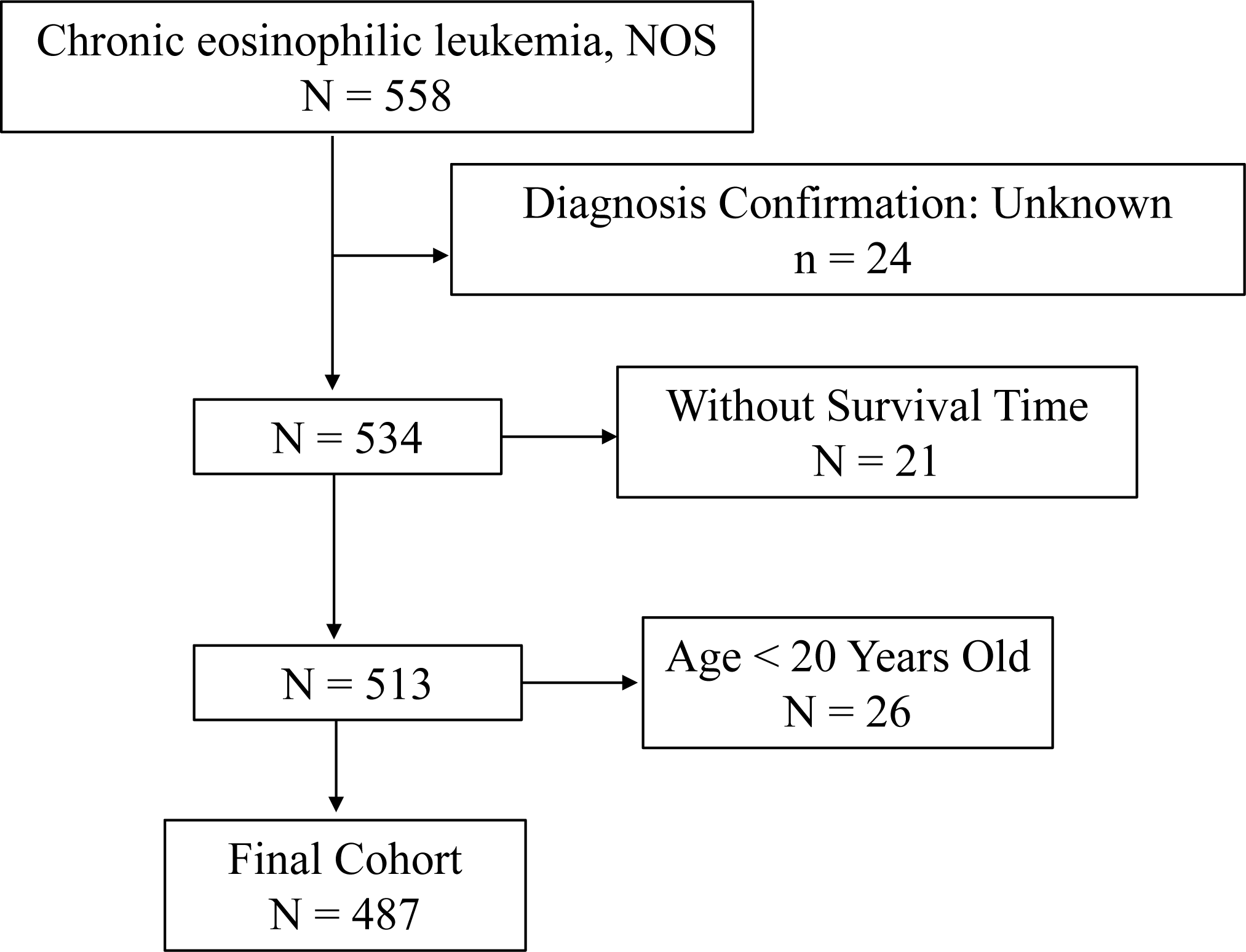
Flow chart of study cohort selection using the SEER database. A flow diagram of selection of patients with CEL-NOS in this study. CEL-NOS, chronic eosinophilic leukemia, not otherwise specified; SEER, Surveillance, Epidemiology, and End Results.

### LASSO Regression and Independent prognostic factors selection

A total of 7 clinical parameters were included in the training set. According to the results of LASSO Cox regression analysis, age, sex, marital status at diagnosis, and sequence were identified for OS risk factors by using the minimum standard value as the criterion (Figure 3). The Cox regression model was further used to screen the prognostic factors. All the four variables passed the preliminary proportional hazards assumption test: age (*P* = 0.746), sex (*P* = 0.253), marital status (*P* = 0.574) and sequence (*P* = 0.720). Univariate Cox regression analysis revealed that age, marital status at diagnosis, and sequence were significantly associated with OS (Table 2). In the multivariate Cox analysis of OS, age, marital status at diagnosis, and sequence were independently and significantly associated with OS (Table 2). Older age (HR 3.74, 95% CI: 2.51-5.60, *P*_J<_J0.001, Table 2), marital status of single (HR 2.44; 95% CI: 1.49-4.00, *P*_J<_J0.001, Table 2), marital status of other (HR 2.08; 95% CI: 1.41-3.10, *P*_J<_J0.001, Table 2), secondary CEL-NOS (HR 1.98; 95% CI: 1.23-3.20, *P*_J= 0.005, Table 2) were significantly associated with worse overall survival. The detailed data was demonstrated in Table 2.

**Figure 3.**
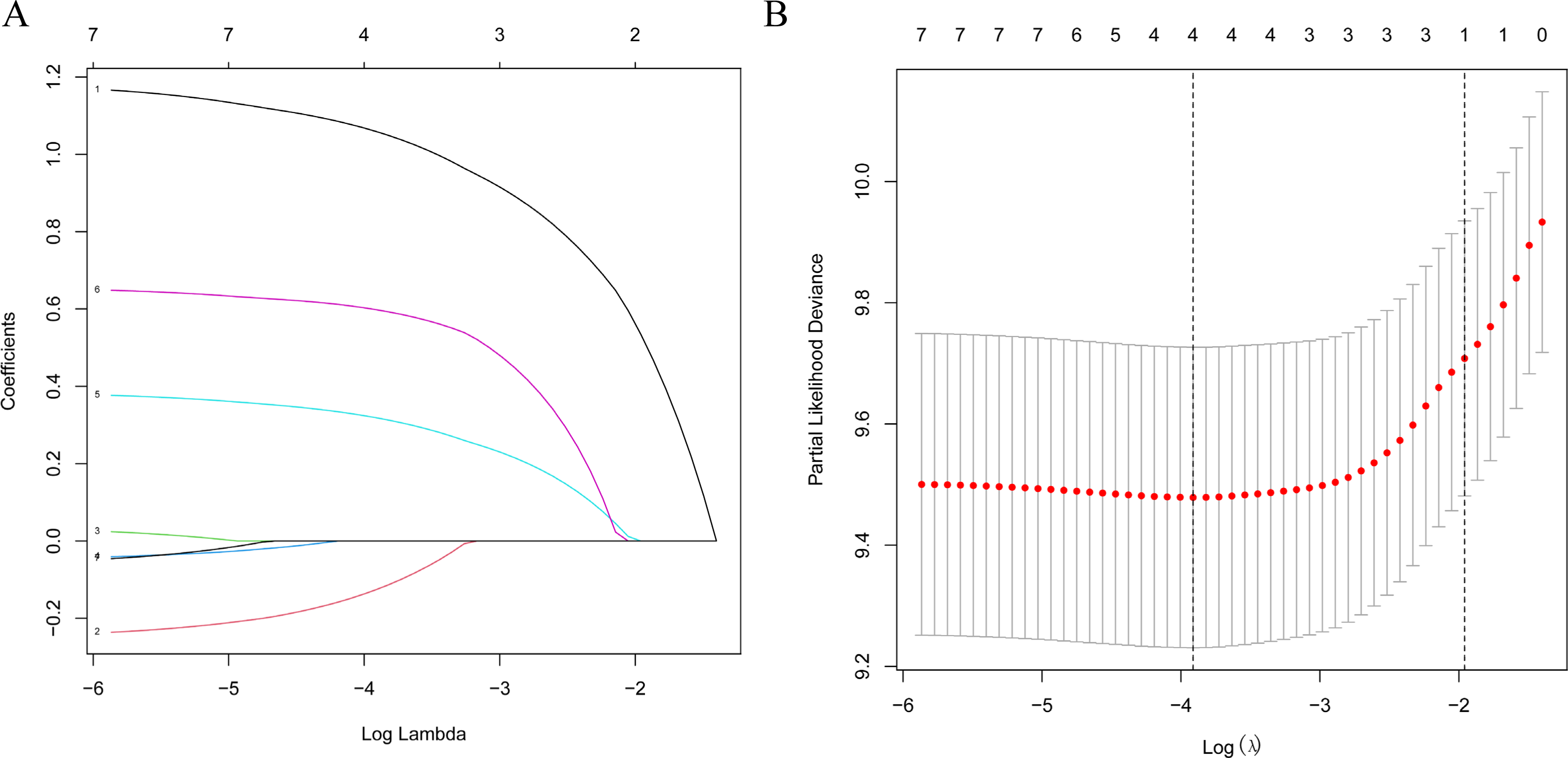
LASSO regression model was used to select characteristic impact factors. (A) LASSO coefficients of 7 features; (B) Selection of tuning parameter (λ) for LASSO model.

### Construction of Prognostic Nomogram

By incorporating the three independent prognostic factors including age, marital status at diagnosis, and sequence, a nomogram was constructed to predict the 3- and 5-year OS probability of CEL-NOS patients (Figure 4). The total points were calculated by integrating scores related to age, marital status, sequence and projected to the bottom scale to predict the OS probability at 3 and 5 years.

**Figure 4.**
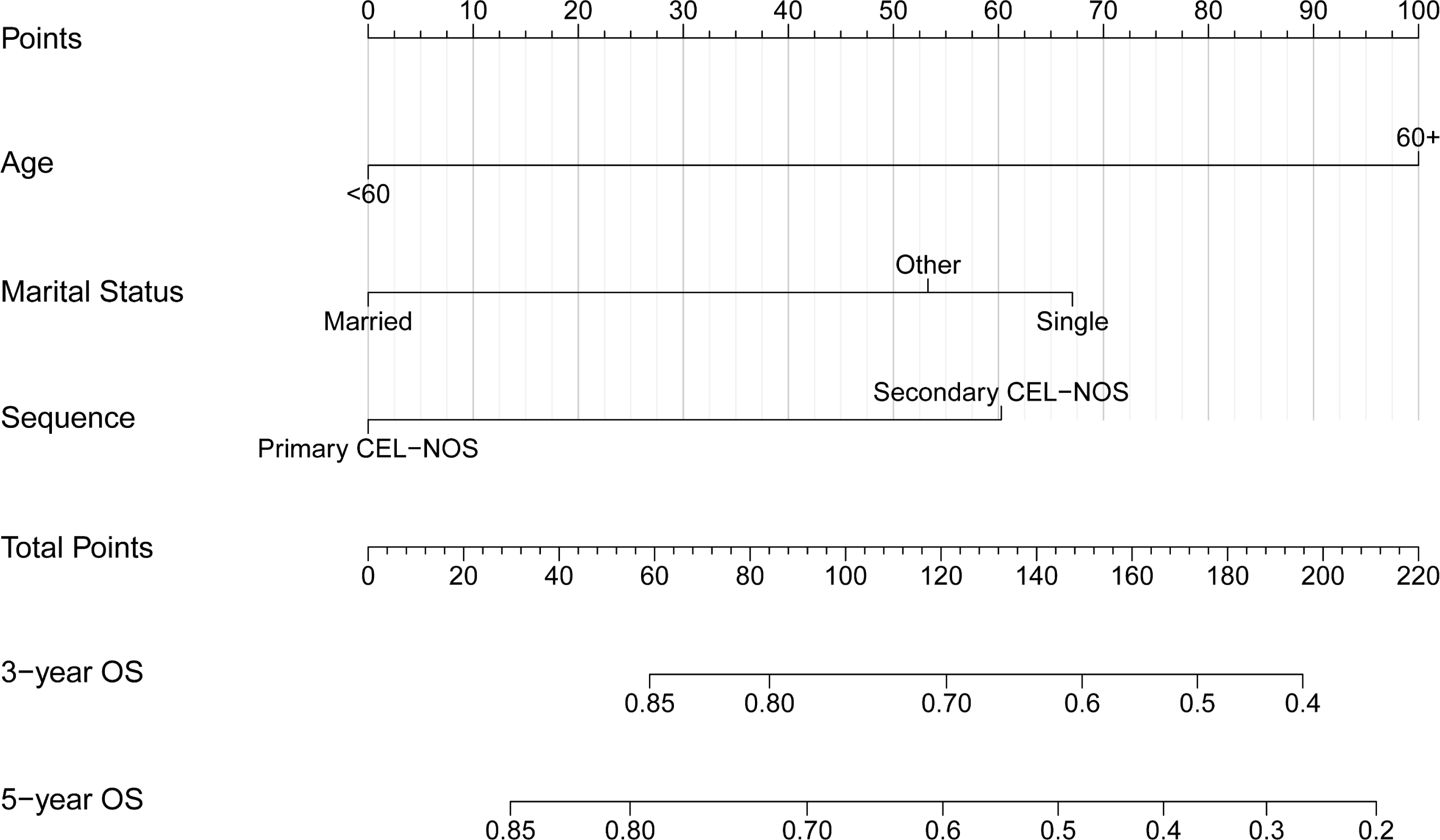
Construction of the prognostic nomogram of CEL-NOS patients based on 3 risk factors. The total points were calculated by integrating scores related to age, marital status, sequence and projected to the bottom scale to predict the overall survival probability at 3 and 5 years. CEL-NOS, chronic eosinophilic leukemia, not otherwise specified.

### Evaluation and Validation of the Nomogram

The calibration curve of the nomogram for the training cohort revealed a close match between the predicted and observed OS probability at the 3- and 5-year intervals (Figure 5A).

**Figure 5.**
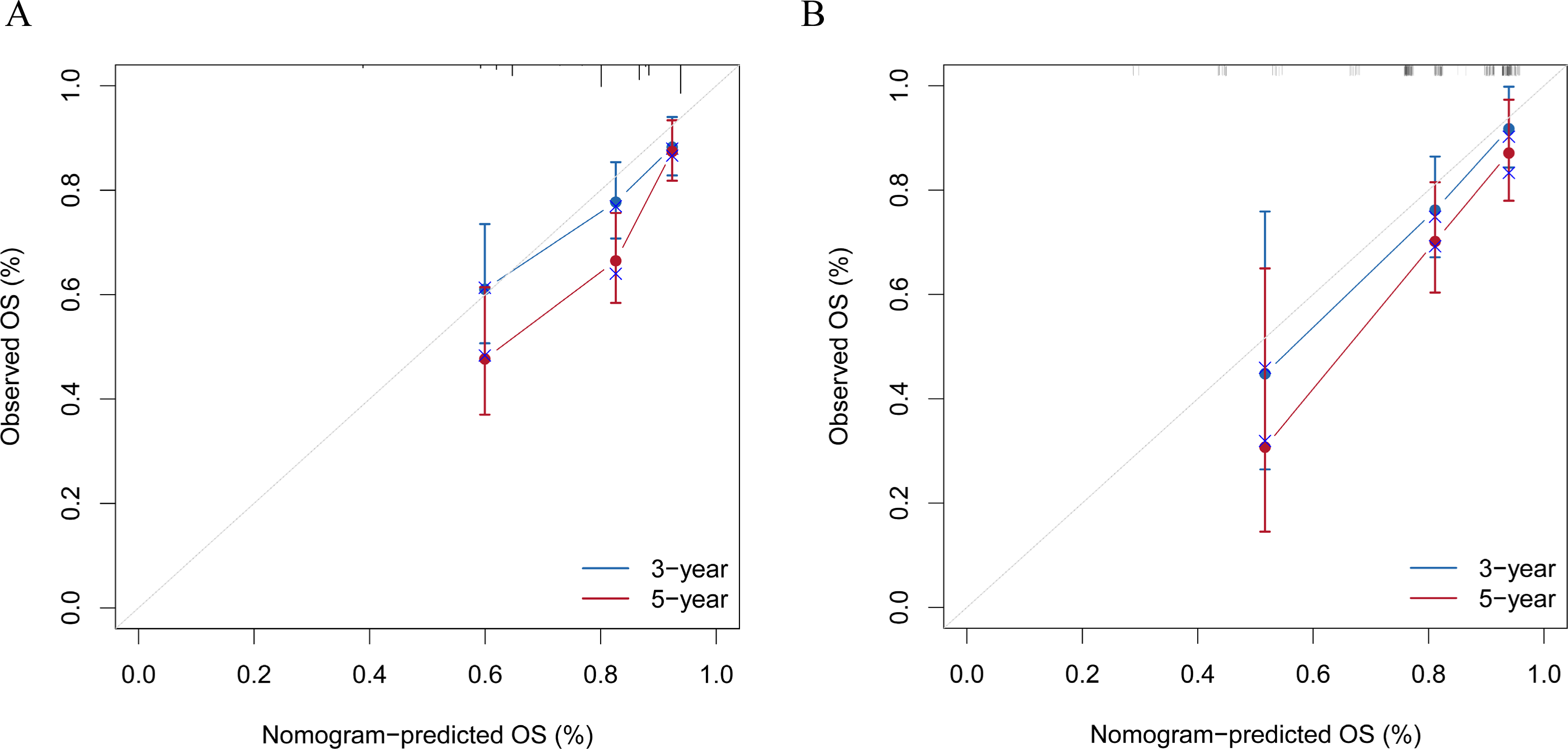
Evaluation of the nomogram of by calibration plot. (A) The calibration curve of the training set for the observed overall survival (OS) probability and predicted OS at 3-year and 5-year. (B) The calibration curve of the validation set at 3-year and 5-year.

Additionally, validation cohort calibration plots at 3- and 5 years also showed good agreement between prediction and actual observation (Figure 5B). Time-dependent ROC analyses showed the accuracy of the nomogram models in predicting 3- and 5-year OS probability in the training set, with AUC values of 0.691 and 0.744, respectively (Figure 6A), and the 3-year and 5-year AUC of the validation set was of 0.696 and 0.708, respectively (Figure 6B). The DCA was employed to evaluate the clinical net benefit of the predictive model. The results showed that the nomogram model has a good net benefit in predicting the 3- and 5-year OS probability both in the training set (Figure 7A, B) and validation set (Figure 7C, D).

**Figure 6.**
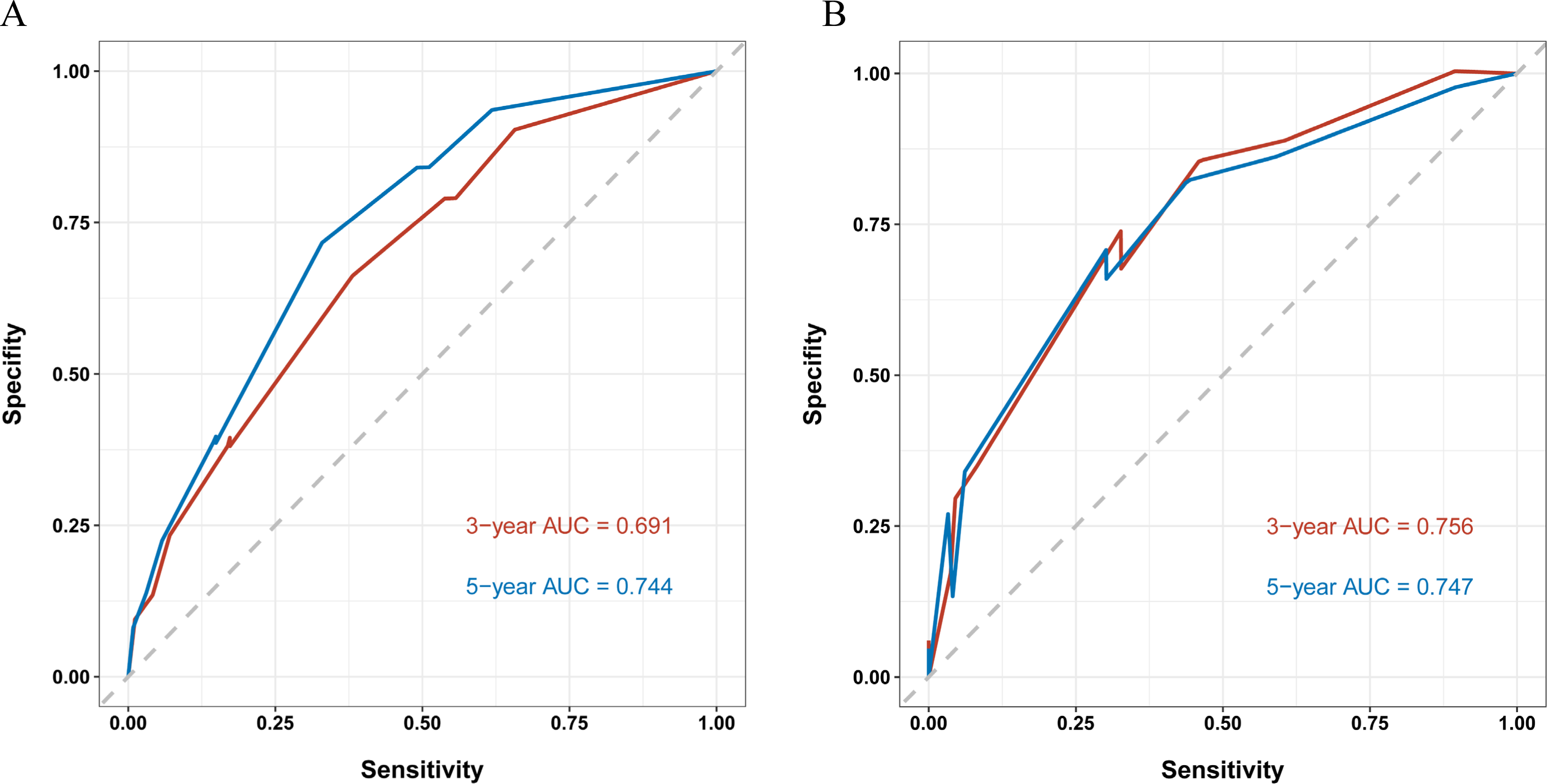
Evaluation of the nomogram of by receiver operating characteristic (ROC) plot. (A) Time-dependent ROC curve analyses of the nomograms for the 3 years and 5 years in the training set. (B) Time-dependent ROC curve analyses of the validation set.

**Figure 7.**
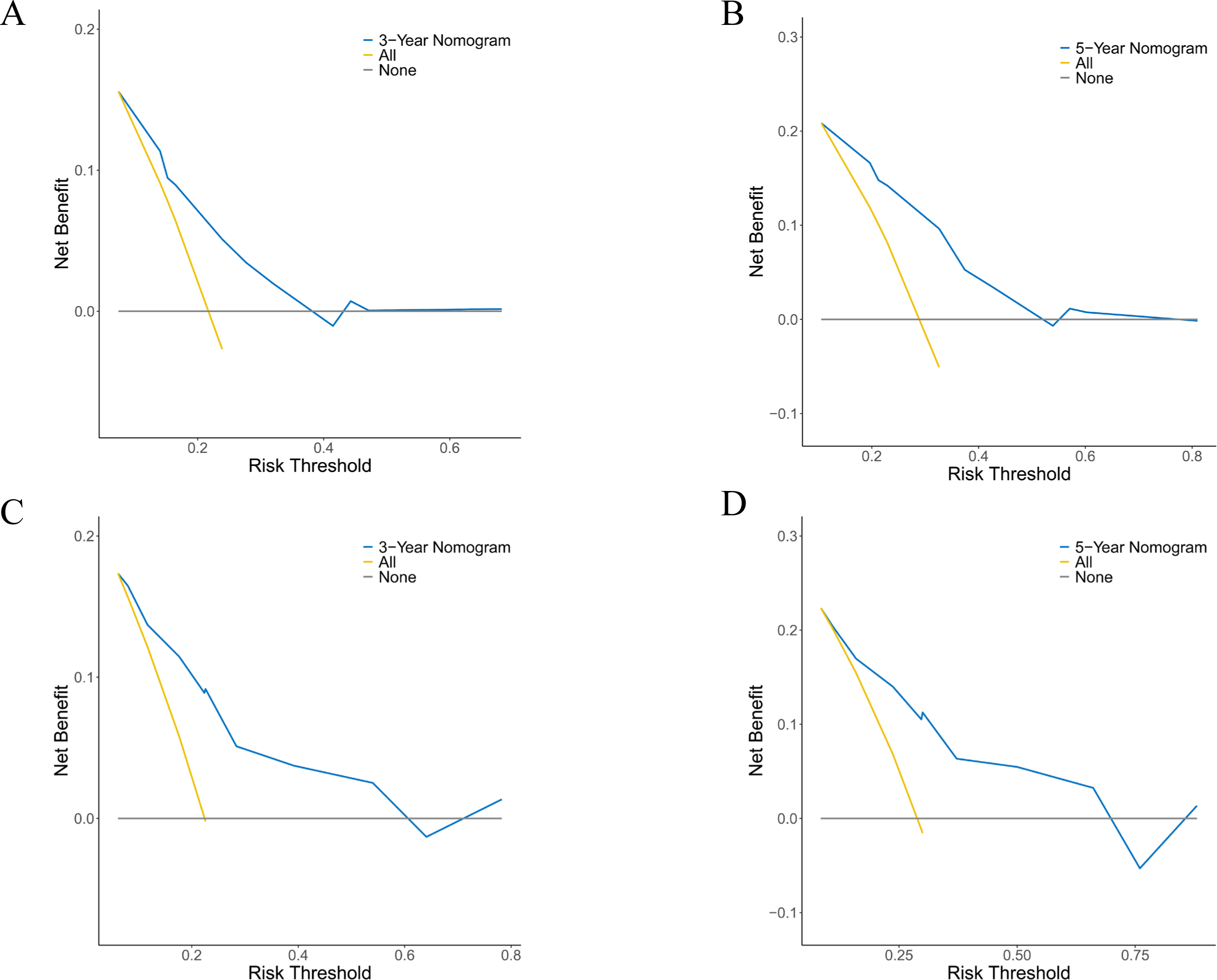
Evaluation of the nomogram of by decision curve analyses. (A, B) The decision curve analyses of the nomogram for the 3 years (A) and 5 years (B) in the training set. (C, D) The decision curve analysis of the nomogram for the 3 years (C) and 5 years (D) in the validation set.

### Survival Analysis Between the Stratified Risk Groups

The score of each variable was generated from the nomogram and the cumulative scores were calculated for all the patients. The entire cohort was stratified into low- and high-risk subgroups according to the median risk score. Kaplan-Meier analysis of OS revealed significant differences between the low- and high -risk groups for both training set (*P*_J<_J0.0001, Figure 8A) and validation set (*P*_J< 0.0001, Figure 8B), which underscores the exceptional capacity of the nomogram for effective risk stratification.

**Figure 8.**
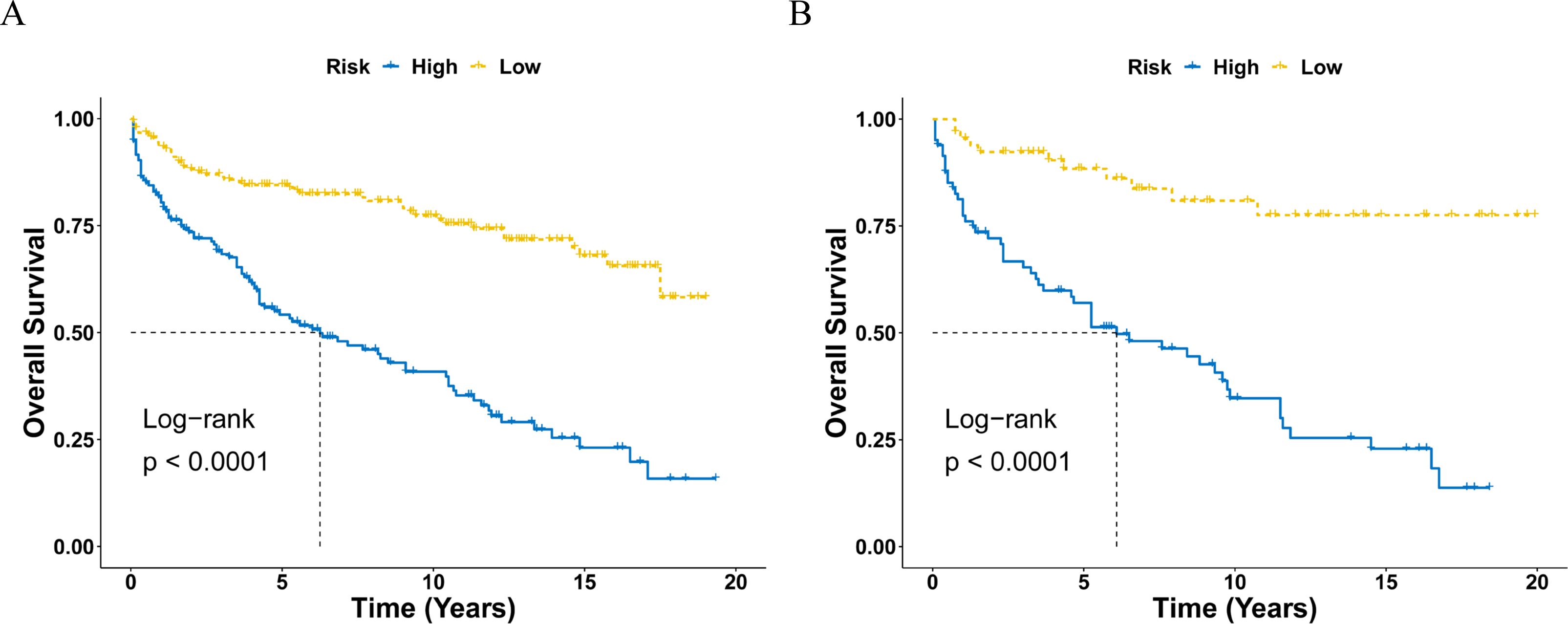
Kaplan-Meier curve of overall survival of CEL-NOS patients stratified by the risk stratification system in the training set (A) and validation set (B). CEL-NOS, chronic eosinophilic leukemia, not otherwise specified.

## Discussion

CEL-NOS represents a rare and intricate hematological disorder characterized by uncontrolled eosinophilic proliferation^8^. Given its rarity, there is limited published literature on CEL-NOS, mostly comprising case reports or small case series, the incidence and clinical characteristics have not been comprehensively studied yet^7,14^. The present study identified 487 CEL-NOS patients from 2001-2020 using the SEER database, representing the largest cohort describing the incidence and clinical characteristics of CEL-NOS patients to date. We also developed and validated a nomogram to predict the 3- and 5-year overall survival probability of CEL-NOS patients based on the screened prognostic factors.

The epidemiology of CEL-NOS remains incompletely characterized due to its rarity and clinical heterogeneity^1,2^. Available studies suggest an estimated incidence rate of approximately 0.036 per 100,000 individuals for all hypereosinophilic syndromes (HES), including CEL-NOS^15^. In this study, analysis of CEL-NOS from the SEER database between 2001 and 2020 revealed a low age-adjusted incidence rate (AIR) of 0.033 per 100,000 person-years. Moreover, it was once reported that CEL-NOS exhibits a male predominance, and the median age at diagnosis was 62 years^7^. In this study, the average age of the patients was 57.0 ± 17.0 years, and the IRR of male-to-female was 1.66 (95% CI, 1.39-1.98), which is in agreement with the previous research.

Furthermore, the current study demonstrated that the incidence rate increased with age, with the IRR of the 60+ age group to the <60 age group being 3.65 (95% CI, 3.07-4.34), which has not been documented in the literature so far. The age distribution of CEL-NOS may reflect the accumulation of genetic and epigenetic alterations that lead to clonal expansion of eosinophils over time^16^. The gender difference of CEL-NOS may be influenced by hormonal factor or genetic factors that affect the susceptibility or exposure to eosinophilic stimuli^17,18^.

A previous study showed that the prognosis of CEL-NOS is poor in a cohort of 10 patients and the median survival was 22.2 months, with 5 patients developing acute transformation after median of 20 months from diagnosis^7^. However, it is extremely difficult to draw any conclusion from this study due to the small sample size. In the current study, to identify the prognostic factors for CEL-NOS, LASSO Cox regression analysis and multivariate Cox regression analysis were performed on a set of clinical variables. The study unveiled that age, marital status at diagnosis, and sequence were independently associated with overall survival. Older age emerged as a significant adverse prognostic factor, with older individuals facing substantially elevated mortality risks (HR 3.74, 95% CI: 2.51-5.60), which is in line with previous studies on MPNs^19^. Marital status was also a significant predictor of survival, with the marital statuses of single and other (divorced, separated, widowed, unmarried or domestic partner) having a worse outcome than married patients. This may reflect the impact of social support and psychological factors on cancer survival^20-22^. Sequence was another important prognostic factor, with secondary CEL-NOS associated with poorer prognosis. This may be due to the presence of other malignancies or comorbidities that affect the treatment response and tolerance^23^. In addition, chemotherapy showed no effect on the OS of CEL-NOS patients in this study, which is in accordance with some study showed that CEL-NOS patients are usually unresponsive to conventional chemotherapy^7^.

Nomograms have been developed and proven to surpass the conventional staging systems in terms of prognostic accuracy for certain types of cancers^24^. Consequently, the integration of nomograms into clinical practice as reliable tools for predicting cancer prognosis has become increasingly prevalent^24,25^. In this study, a prognostic nomogram was constructed to predict the 3- and 5-year overall survival probability of CEL-NOS patients based on those three independent prognostic factors: age, marital status at diagnosis, and sequence. The nomogram demonstrated commendable calibration and discriminative performance in both the training and validation cohorts, indicating satisfactory accuracy and consistency of the nomogram. This study is the first to develop and validate a clinical prognostic model for CEL-NOS patients. To the best of our knowledge, this is also the largest study ever conducted.

Effective risk stratification is integral to tailoring treatment strategies and optimizing patient outcomes^26^. Utilizing the nomogram, CEL-NOS patients were stratified into low- and high-risk groups based on individual risk scores. Kaplan-Meier analysis of OS revealed substantial distinctions between these risk cohorts, underscoring the nomogram’s efficacy in risk stratification. This empowers clinicians to identify patients who may benefit from more aggressive therapeutic interventions or intensified surveillance, ultimately contributing to improved patient care and outcomes.

However, there are some limitations with this study. Firstly, the SEER registry did not document other potential prognostic factors that may have a significant impact on CEL-NOS patient outcomes, such as genetic mutation, performance status, LDH level, immunophenotypic features, family history and alcohol/smoking consumption history. Secondly, detailed information about therapy was not recorded in the SEER database, making it impossible to analyze the effect of different treatment regimens. Thirdly, this is a retrospective study, which means that there may be unavoidable potential biases such as selection bias. Finally, while the nomograms of CEL-NOS were constructed and verified using the same database, they were not further validated using another independent dataset. Thus, although this study provided important insights on CEL-NOS due to the rarity and lack of large-scale multicenter prospective study of this disease, the results should still be interpreted with caution.

## Conclusions

In conclusion, this study provides novel insights into the epidemiology and prognosis of CEL-NOS in the US population using the SEER database. CEL-NOS is a very rare disease with a variable clinical presentation and outcome. Age, marital status at diagnosis, and sequence were identified as independent prognostic factors for overall survival, culminating in the development of a prognostic nomogram to predict the 3- and 5-year overall survival probability of CEL-NOS patients. This nomogram may help clinicians provide personalized treatment and clinical decisions for CEL-NOS patients. To our knowledge, this study represents the largest population-based cohort investigating the epidemiology and survival outcome of CEL patients. However, more clinical research is needed to validate our findings.

## Data Availability Statement

The data analyzed in this study are from the SEER database (https://seer.cancer.gov/) that are available to the public.

## Acknowledgments

The interpretation of the data is the sole responsibility of the author(s). The author(s) acknowledge the efforts of the National Cancer Institute and the Surveillance, Epidemiology, and End Results (SEER) Program tumor registries in the creation of the SEER database.

## Abbreviations

CEL: Chronic eosinophilic leukemia
Chemo: chemotherapy
CI: confidence interval
COD: cause of death
HR: hazard ration
OS: overall survival
SEER: Surveillance, Epidemiology, and End Results
NOS: not otherwise specified

## References

1. Arber DA, Orazi A, Hasserjian R, et al. The 2016 revision to the World Health Organization classification of myeloid neoplasms and acute leukemia. Blood. 2016/05/19/ 2016;127(20):2391-2405. 10.1182/blood-2016-03-643544

2. Shomali W, Gotlib J. World Health Organization-defined eosinophilic disorders: 2019 update on diagnosis, risk stratification, and management. American Journal of Hematology. 2019;94(10):1149–1167. 10.1002/ajh.25617

3. Pardanani A, D’Souza A, Knudson RA, Hanson CA, Ketterling RP, Tefferi A. Long-term follow-up of FIP1L1-PDGFRA-mutated patients with eosinophilia: survival and clinical outcome. Leukemia. 2012/11/01 2012;26(11):2439-2441. doi:10.1038/leu.2012.162

4. Ogbogu PU, Bochner BS, Butterfield JH, et al. Hypereosinophilic syndrome: A multicenter, retrospective analysis of clinical characteristics and response to therapy. Journal of Allergy and Clinical Immunology. 2009;124(6):1319–1325.e3. doi:10.1016/j.jaci.2009.09.022

5. Wang SA, Tam W, Tsai AG, et al. Targeted next-generation sequencing identifies a subset of idiopathic hypereosinophilic syndrome with features similar to chronic eosinophilic leukemia, not otherwise specified. Modern Pathology. 2016/08/01/ 2016;29(8):854-864. 10.1038/modpathol.2016.75

6. Sa AW, Robert PH, Wayne T, et al. Bone marrow morphology is a strong discriminator between chronic eosinophilic leukemia, not otherwise specified and reactive idiopathic hypereosinophilic syndrome. Haematologica. 08/01 2017;102(8):1352-1360. doi:10.3324/haematol.2017.165340

7. Helbig G, Soja A, Bartkowska-Chrobok A, Kyrcz-Krzemień S. Chronic eosinophilic leukemia-not otherwise specified has a poor prognosis with unresponsiveness to conventional treatment and high risk of acute transformation. American Journal of Hematology. 2012/06/01 2012;87(6):643-645. 10.1002/ajh.23193

8. Gotlib J. World Health Organization-defined eosinophilic disorders: 2017 update on diagnosis, risk stratification, and management. American Journal of Hematology. 2017/11/01 2017;92(11):1243-1259. 10.1002/ajh.24880

9. Helbig G, Moskwa A, Hus M, et al. Durable remission after treatment with very low doses of imatinib for FIP1L1-PDGFRα-positive chronic eosinophilic leukaemia. Cancer Chemotherapy and Pharmacology. 2011/04/01 2011;67(4):967-969. doi:10.1007/s00280-011-1582-3

10. Metzgeroth G, Walz C, Erben P, et al. Safety and efficacy of imatinib in chronic eosinophilic leukaemia and hypereosinophilic syndrome – a phase-II study. British Journal of Haematology. 2008/12/01 2008;143(5):707-715. 10.1111/j.1365-2141.2008.07294.x

11. Yamada O, Kitahara K, Imamura K, Ozasa H, Okada M, Mizoguchi H. Clinical and cytogenetic remission induced by interferon-α in a patient with chronic eosinophilic leukemia associated with a unique t(3;9;5) translocation. American Journal of Hematology. 1998/06/01 1998;58(2):137-141. 10.1002/(SICI)1096-8652(199806)58:2<137::AID-AJH9>3.0.CO;2-T

12. Basara N, Markova J, Schmetzer B, et al. Chronic Eosinophilic Leukemia: Successful Treatment with an Unrelated Bone Marrow Transplantation. Leukemia & Lymphoma. 1998/01/01 1998;32(1-2):189-193. doi:10.3109/10428199809059261

13. H. Dunphy C. Chronic Eosinophilic Leukemia, Not Otherwise Specified (CEL, NOS). Current Cancer Therapy Reviews. 2012;8(1):30-34. doi:10.2174/157339412799462512

14. Morsia E, Reichard K, Pardanani A, Tefferi A, Gangat N. WHO defined chronic eosinophilic leukemia, not otherwise specified (CEL, NOS): A contemporary series from the Mayo Clinic. American Journal of Hematology. 2020/07/01 2020;95(7):E172-E174. 10.1002/ajh.25811

15. Crane MM, Chang CM, Kobayashi MG, Weller PF. Incidence of myeloproliferative hypereosinophilic syndrome in the United States and an estimate of all hypereosinophilic syndrome incidence. Journal of Allergy and Clinical Immunology. 2010;126(1):179–181. doi:10.1016/j.jaci.2010.03.035

16. Aldoss I, Forman SJ, Pullarkat V. Acute Lymphoblastic Leukemia in the Older Adult. Journal of Oncology Practice. 2019/02/01 2019;15(2):67-75. doi:10.1200/JOP.18.00271

17. Garner WB, Smith BD, Shabason JE, et al. Predicting future cancer incidence by age and gender. Journal of Clinical Oncology. 2019/05/20 2019;37(15_suppl):1559-1559. doi:10.1200/JCO.2019.37.15_suppl.1559

18. Ornos ED, Cando LF, Catral CD, et al. Molecular basis of sex differences in cancer: Perspective from Asia. iScience. 2023/07/21/ 2023;26(7):107101. 10.1016/j.isci.2023.107101

19. Hultcrantz M, Kristinsson SY, Andersson TML, et al. Patterns of Survival Among Patients With Myeloproliferative Neoplasms Diagnosed in Sweden From 1973 to 2008: A Population-Based Study. Journal of Clinical Oncology. 2012/08/20 2012;30(24):2995-3001. doi:10.1200/JCO.2012.42.1925

20. Aizer AA, Chen M-H, McCarthy EP, et al. Marital Status and Survival in Patients With Cancer. Journal of Clinical Oncology. 2013;31(31):3869–3876. doi:10.1200/jco.2013.49.6489

21. Ding Z, Yu D, Li H, Ding Y. Effects of marital status on overall and cancer-specific survival in laryngeal cancer patients: a population-based study. Scientific Reports. 2021/01/12 2021;11(1):723. doi:10.1038/s41598-020-80698-z

22. Kravdal Ø. The impact of marital status on cancer survival. Social Science & Medicine. 2001/02/01/ 2001;52(3):357-368. doi:10.1016/S0277-9536(00)00139-8

23. Keegan THM, Bleyer A, Rosenberg AS, Li Q, Goldfarb M. Second Primary Malignant Neoplasms and Survival in Adolescent and Young Adult Cancer Survivors. JAMA Oncology. 2017;3(11):1554–1557. doi:10.1001/jamaoncol.2017.0465

24. Iasonos A, Schrag D, Raj GV, Panageas KS. How To Build and Interpret a Nomogram for Cancer Prognosis. Journal of Clinical Oncology. 2008/03/10 2008;26(8):1364-1370. doi:10.1200/JCO.2007.12.9791

25. Balachandran VP, Gonen M, Smith JJ, DeMatteo RP. Nomograms in oncology: more than meets the eye. The Lancet Oncology. 2015;16(4):e173–e180. doi:10.1016/S1470-2045(14)71116-7

26. Watson EK, Rose PW, Neal RD, et al. Personalised cancer follow-up: risk stratification, needs assessment or both? British Journal of Cancer. 2012/01/01 2012;106(1):1-5. doi:10.1038/bjc.2011.535

