## Supplementary material for "Epidemiology and Nomogram Development for Chronic Eosinophilic Leukemia, Not Otherwise Specified (CEL-NOS): Insights from the SEER Database": Table 1

**Table 1** Comparison of baseline characteristics of patients with CEL-NOS between the training set and validation set from SEER database.

| **Characteristics** | **Total (N=487)** | **Training set (N=339)** | **Validation set (N=148)** | ***P*-value** |
| --- | --- | --- | --- | --- |
| **Sex** |  |  |  |  |
| Male | 298 (61.2%) | 208 (61.4%) | 90 (60.8%) | 0.990 |
| Female | 189 (38.8%) | 131 (38.6%) | 58 (39.2%) |  |
| **Age** |  |  |  |  |
| <60 | 260 (53.4%) | 191 (56.3%) | 69 (46.6%) | 0.060 |
| 60+ | 227 (46.6%) | 148 (43.7%) | 79 (53.4%) |  |
| **Race** |  |  |  |  |
| White | 360 (73.9%) | 245 (72.3%) | 115 (77.7%) | 0.452 |
| African American | 76 (15.6%) | 56 (16.5%) | 20 (13.5%) |  |
| Other^1^ | 51 (10.5%) | 38 (11.2%) | 13 (8.8%) |  |
| **Diagnosis Year** |  |  |  |  |
| 2001-2005 | 136 (27.9%) | 104 (30.7%) | 32 (21.6%) | 0.091 |
| 2006-2010 | 134 (27.5%) | 96 (28.3%) | 38 (25.7%) |  |
| 2011-2015 | 113 (23.2%) | 72 (21.2%) | 41 (27.7%) |  |
| 2016-2020 | 104 (21.4%) | 67 (19.8%) | 37 (25.0%) |  |
| **Sequence** |  |  |  |  |
| Primary CEL-NOS^2^ | 437 (89.7%) | 307 (90.6%) | 130 (87.8%) | 0.454 |
| Secondary CEL-NOS^3^ | 50 (10.3%) | 32 (9.4%) | 18 (12.2%) |  |
| **Marital Status** |  |  |  |  |
| Married | 279 (57.3%) | 193 (56.9%) | 86 (58.1%) | 0.905 |
| Single^4^ | 88 (18.1%) | 63 (18.6%) | 25 (16.9%) |  |
| Other^5^ | 120 (24.6%) | 83 (24.5%) | 37 (25.0%) |  |
| **Chemo** |  |  |  |  |
| Yes | 201 (41.3%) | 146 (43.1%) | 55 (37.2%) | 0.264 |
| No/Unknown | 286 (58.7%) | 193 (56.9%) | 93 (62.8%) |  |
| **COD** |  |  |  |  |
| Alive | 284 (58.3%) | 200 (59.0%) | 84 (56.8%) | 0.960 |
| CEL-NOS^6^ | 42 (8.6%) | 29 (8.6%) | 13 (8.8%) |  |
| Heart Diseases | 45 (9.2%) | 30 (8.8%) | 15 (10.1%) |  |
| Other cause^7^ | 116 (23.8%) | 80 (23.6%) | 36 (24.3%) |  |

^1^ Races including Asian/Pacific Islander, American Indian/Alaska Native and “unknown”.

^2^ Sequence number of “primary only” and “1st of 2 or more primaries”, indicating CEL-NOS was the primary malignancy.

^3^ Sequence number of “2nd of 2 or more primaries”, “3rd of 3 or more primaries” and “4th of 4 or more primaries”, indicating CEL-NOS was secondary to other primary malignancies.

^4^ Marital status at diagnosis was single (never married).

^5^ Marital statuses of divorced, widowed, separated, unmarried or domestic partner and unknown at diagnosis.

^6^ Death attributable to CEL-NOS.

^7^ Dead of other causes such as diabetes mellitus, cerebrovascular diseases, septicemia, and so on.

Abbreviations: CEL-NOS, chronic eosinophilic leukemia, not otherwise specified; Chemo, chemotherapy; COD, cause of death.
