## Supplementary material for "Epidemiology and Nomogram Development for Chronic Eosinophilic Leukemia, Not Otherwise Specified (CEL-NOS): Insights from the SEER Database": Table 2

**Table 3** Univariable and multivariable cox regression analysis for overall survival of patients with CEL-NOS.

| **Parameters** | **Univariable** | | | **Multivariable** | | |
| --- | --- | --- | --- | --- | --- | --- |
|  | **HR** | **95% CI** | ***P*-value** | **HR** | **95% CI** | ***P*-value** |
| **Age** |  |  |  |  |  |  |
| <60 | *ref* |  |  | *ref* |  |  |
| 60+ | 3.50 | [2.46, 4.98] | < 0.001 | 3.74 | [2.51, 5.60] | < 0.001 |
| **Sex** |  |  |  |  |  |  |
| Male | *ref* |  |  | *ref* |  |  |
| Female | 0.97 | [0.69, 1.37] | 0.900 | 0.76 | [0.53, 1.10] | 0.146 |
| **Marital Status** |  |  |  |  |  |  |
| Married | *ref* |  |  | *ref* |  |  |
| Single^1^ | 1.38 | [0.87, 2.17] | 0.200 | 2.44 | [1.49, 4.00] | < 0.001 |
| Other^2^ | 2.20 | [1.51, 3.21] | < 0.001 | 2.08 | [1.41, 3.10] | < 0.001 |
| **Sequence** |  |  |  |  |  |  |
| Primary^3^ | ref |  |  | ref |  |  |
| Secondary^4^ | 2.87 | [1.83, 4.50] | < 0.001 | 1.98 | [1.23, 3.20] | 0.005 |

^1^ Marital status of single (never married) at diagnosis.

^2^ Marital statuses of divorced, widowed, separated, unmarried or domestic partner and unknown at diagnosis.

^3^ Consisted of “one primary only” and “1st of 2 or more primaries”.

^4^ Consisted of “2nd of 2 or more primaries”, “3rd of 3 or more primaries” and “4th of 4 or more primaries”, indicating CEL-NOS was secondary to other primary malignancies.

Abbreviations: CEL-NOS, chronic eosinophilic leukemia, not otherwise specified; Chemo, chemotherapy; CI, confidence interval; HR, hazard ration; OS, overall survival.
